# Multimodal Radiogenomic Machine Learning for Biochemical Recurrence Prediction Following Radical Prostatectomy Using PSMA-PET, mpMRI, and the Decipher Genomic Classifier

**DOI:** 10.64898/2026.08.05.26359806

**Authors:** Ruchika Reddy Chimmula, Courtney Yong, Harrison Louis Love, Rakesh Shiradkar, Jordan Holmes, Varsha Nair, Mark Tann, Clint Bahler, Oluwaseyi Oderinde

## Abstract

**Background:** Biochemical recurrence (BCR) occurs in up to 40% of men following radical prostatectomy (RP). Current risk models rely primarily on clinicopathologic variables and may not fully capture the biological heterogeneity associated with recurrence. The Decipher Genomic Classifier (DGC), prostate-specific membrane antigen positron emission tomography (PSMA-PET), and multiparametric magnetic resonance imaging (mpMRI) provide complementary prognostic information that may improve prediction.

**Objective:** To develop and evaluate machine learning (ML) models integrating DGC, PSMA-PET, and mpMRI for preoperative prediction of BCR following RP.

**Methods:** This retrospective study included patients with available preoperative DGC, PSMA-PET, mpMRI, and clinicopathologic data. Logistic regression (LR), random forest (RF), and XGBoost models were developed using single- and multimodality feature combinations. Early- and intermediate-fusion strategies were evaluated. Performance was assessed using an area under the receiver operating characteristic curve (AUC) and accuracy. Clinical utility was evaluated using decision curve analysis.

**Results:** XGBoost consistently outperformed LR and RF. DGC achieved the highest single-modality performance (AUC 0.94, accuracy 86.7%). Among multimodal models, DGC combined with PSMA-PET using intermediate fusion achieved the best overall performance (AUC 0.93, accuracy 87.0%). Addition of mpMRI reduced performance (AUC 0.85, accuracy 83.0%). Decision curve analysis demonstrated positive net benefit across clinically relevant thresholds.

**Conclusion:** XGBoost-based multimodal fusion improved preoperative BCR prediction following RP. DGC was the strongest individual predictor, while integration with PSMA-PET provided the best overall performance, supporting the potential of radiogenomic ML models for personalized risk stratification.

## 1.0 Introduction

Prostate cancer (PCa) is the mostly diagnosed non-cutaneous malignancy among men and remains a leading cause of cancer-related mortality worldwide [1]. Despite advances in diagnosis and treatment, biochemical recurrence (BCR), defined as a rise in prostate-specific antigen (PSA) following radical prostatectomy (RP), occurs in up to 40% of patients within 10 years of surgery [2,3]. Although BCR does not always lead to clinical progression, it is associated with an increased risk of metastasis and prostate cancer-specific mortality, making accurate preoperative risk stratification an important clinical objective [4–6].

Current risk stratification tools, including the CAPRA score, D’Amico classification, and Kattan nomogram, rely primarily on clinical and pathological variables such as PSA, Gleason grade, and biopsy findings [7–9]. While these models provide useful prognostic information, they do not fully capture the biological heterogeneity of prostate cancer, and patients with similar clinicopathologic characteristics may experience markedly different outcomes.

Several biomarkers have shown promises for improving recurrence prediction. Multiparametric magnetic resonance imaging (mpMRI) offers superior soft tissue contrast, enabling precise anatomical information regarding tumor extent and local invasion, while radiomic approaches have demonstrated the prognostic value of quantitative MRI-derived features [10–12]. Prostate-specific membrane antigen positron emission tomography (PSMA-PET) offers sensitive assessment of biologically active disease, and biomarkers such as SUVmax, tumor volume, and nodal involvement have been associated with BCR risk [13,14]. In parallel, the 22-gene Decipher Genomic Classifier (DGC) has emerged as a validated molecular prognostic tool associated with recurrence, metastasis, and prostate cancer-specific mortality [15–17].

These modalities capture complementary aspects of tumor biology, including molecular aggressiveness, disease burden, and anatomical extent [18]. However, integrating heterogeneous genomic and imaging data remains challenging. Machine learning (ML) methods are well suited for modeling complex relationships within multimodal datasets, and gradient-boosted decision tree algorithms such as XGBoost have demonstrated strong predictive performance in oncology applications [19–21]. Furthermore, multimodal learning studies suggest that intermediate fusion strategies may preserve modality-specific information more effectively than conventional early-fusion approaches.

Therefore, we developed and evaluated machine learning models integrating DGC, PSMA-PET, and mpMRI biomarkers for preoperative prediction of BCR following RP. We compared logistic regression, random forest, and XGBoost models across multiple fusion strategies and modality combinations, assessed clinical utility using decision curve analysis, and translated the best-performing model into an interpretable nomogram for individualized risk estimation.

## 2.0. Materials and Methods

### 2.1. Patient Information/Data Collection

This retrospective study was approved by the Institutional Review Board at Indiana University Hospital (Protocol #13892). A total of 108 prostate cancer patients who underwent radical prostatectomy were included, with approximately 32% experiencing biochemical recurrence (BCR) within 3 years. Clinical variables, including prostate-specific antigen density (PSAD), percentage of positive biopsy cores, and Gleason grade, were obtained from medical records. Additional biomarkers were collected from multiparametric MRI (mpMRI), ^68Ga-PSMA-11 PET imaging, and Decipher Genomic Classifier (DGC) reports. Patient characteristics are summarized in Table 1.

**Table 1:**
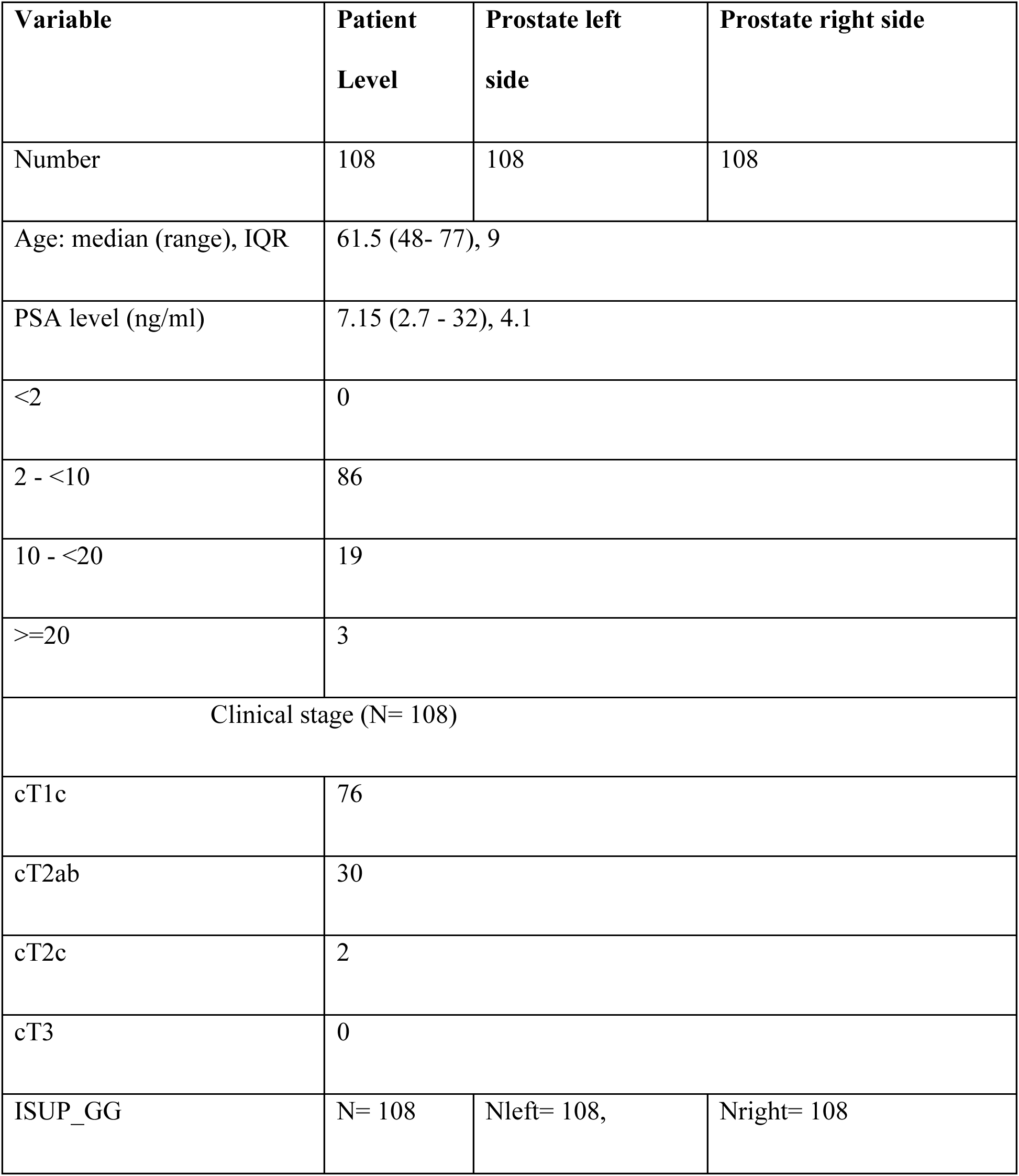
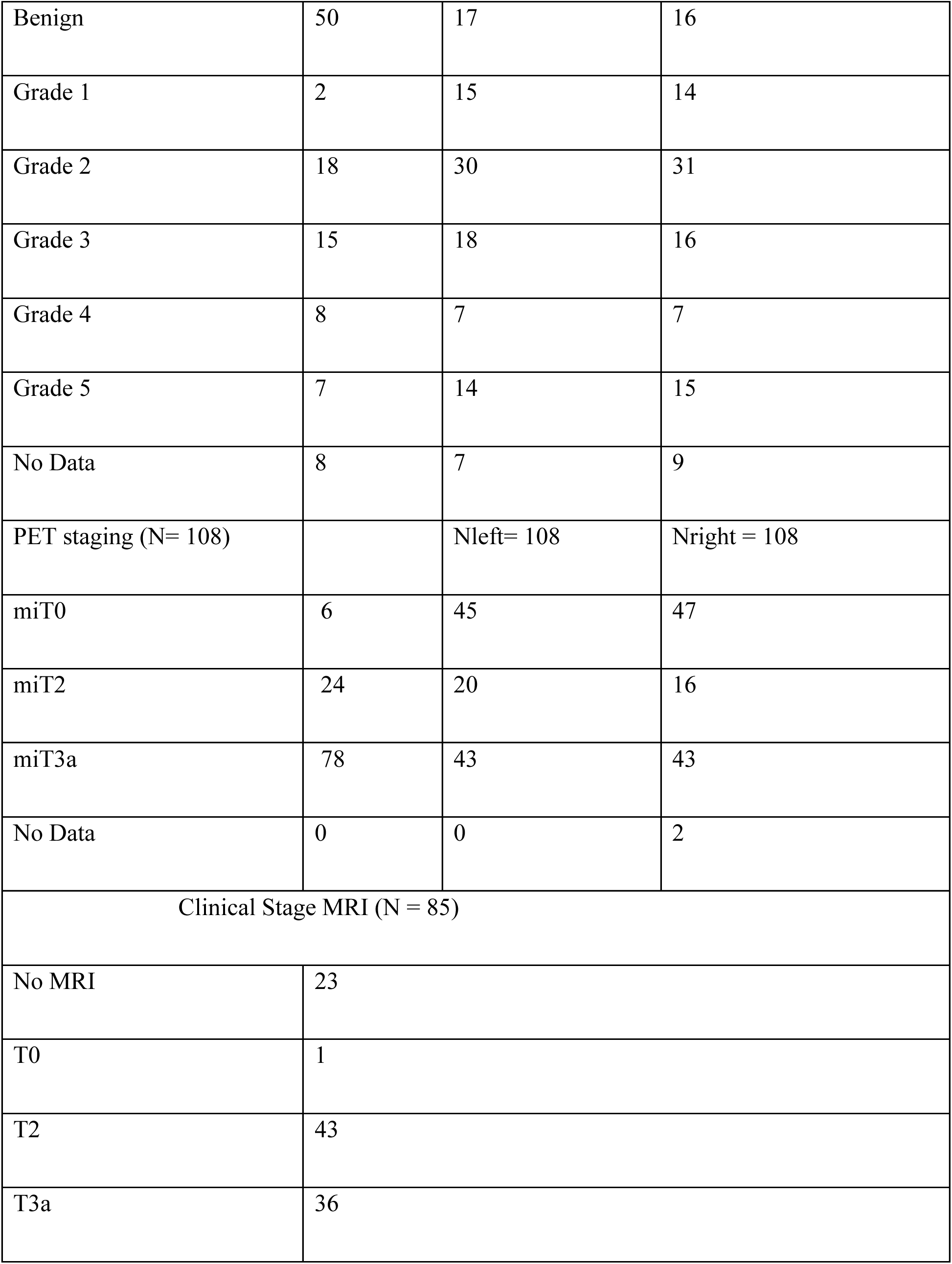
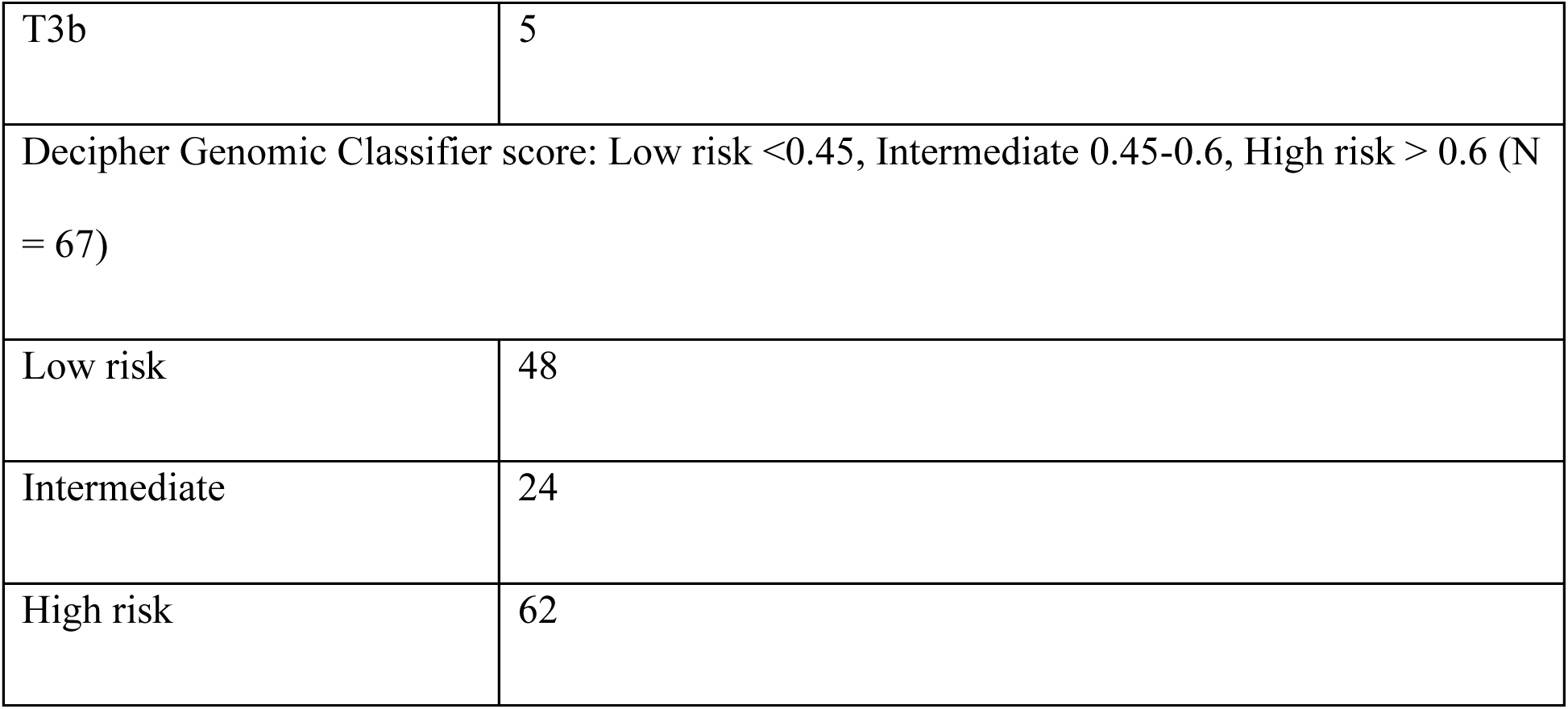
Patients’ Characteristics.

| Variable | Patient Level | Prostate left side | Prostate right side |
| --- | --- | --- | --- |
| Number | 108 | 108 | 108 |
| Age: median (range), IQR | 61.5 (48- 77), 9 |  |  |
| PSA level (ng/ml) | 7.15 (2.7 - 32), 4.1 |  |  |
| <2 | 0 |  |  |
| 2 - <10 | 86 |  |  |
| 10 - <20 | 19 |  |  |
| >=20 | 3 |  |  |
| Clinical stage (N= 108) |  |  |  |
| cT1c | 76 |  |  |
| cT2ab | 30 |  |  |
| cT2c | 2 |  |  |
| cT3 | 0 |  |  |
| ISUP_GG | N= 108 | Nleft= 108, | Nright= 108 |
| Benign | 50 | 17 | 16 |
| Grade 1 | 2 | 15 | 14 |
| Grade 2 | 18 | 30 | 31 |
| Grade 3 | 15 | 18 | 16 |
| Grade 4 | 8 | 7 | 7 |
| Grade 5 | 7 | 14 | 15 |
| No Data | 8 | 7 | 9 |
| PET staging (N= 108) |  | Nleft= 108 | Nright = 108 |
| miT0 | 6 | 45 | 47 |
| miT2 | 24 | 20 | 16 |
| miT3a | 78 | 43 | 43 |
| No Data | 0 | 0 | 2 |
| Clinical Stage MRI (N = 85) |  |  |  |
| No MRI | 23 |  |  |
| T0 | 1 |  |  |
| T2 | 43 |  |  |
| T3a | 36 |  |  |

|  |  |
| --- | --- |
| T3b | 5 |
| Decipher Genomic Classifier score: Low risk <0.45, Intermediate 0.45-0.6, High risk > 0.6 (N = 67) |  |
| Low risk | 48 |
| Intermediate | 24 |
| High risk | 62 |

### 2.2 Imaging and Genomic Biomarkers

Patients underwent standard-of-care mpMRI on 3T scanners, including T2-weighted imaging, diffusion-weighted imaging, and dynamic contrast-enhanced imaging. Tumor PI-RADS scores and lesion diameters were reviewed by an experienced radiologist as previously described ^18^.

Preoperative ^68^Ga-PSMA-11 PET imaging was performed according to institutional protocols. PET scans were interpreted by a dual-board-certified radiologist and nuclear medicine physician, and quantitative imaging biomarkers, including standardized uptake values (SUVs), were recorded.

DGC testing was performed in a CLIA-certified laboratory (Veracyte, San Diego, CA). The Decipher report included the genomic risk score and expression profiles associated with tumor differentiation, proliferation, immune response, metabolism, and other biological pathways. All MRI and PSMA-PET examinations were performed according to institutional clinical protocols. Imaging biomarkers used in this study were extracted from clinically interpreted imaging studies performed by board-certified radiologists. Because the study utilized routinely acquired clinical imaging data, no additional image preprocessing or post hoc quality control procedures were performed. Biopsy procedures were performed as part of routine clinical care. Depending on clinical indication and imaging findings, patients underwent systematic biopsy, MRI-targeted biopsy, or a combination of both according to institutional practice. Biochemical recurrence (BCR) was defined as a postoperative prostate-specific antigen (PSA) level ≥0.2 ng/mL confirmed by a second consecutive PSA measurement, consistent with the American Urological Association/American Society for Radiation Oncology/Society of Urologic Oncology guideline

### 2.3 Data Preprocessing and Feature Selection

Missing data was managed via listwise deletion. Categorical variables were properly encoded to ensure model compatibility, while continuous variables were standardized prior to model training. Exploratory data analysis and correlation assessment were performed to identify relationships among clinical, imaging, and genomic variables and to guide feature selection (Figure 1).

**Figure 1:**
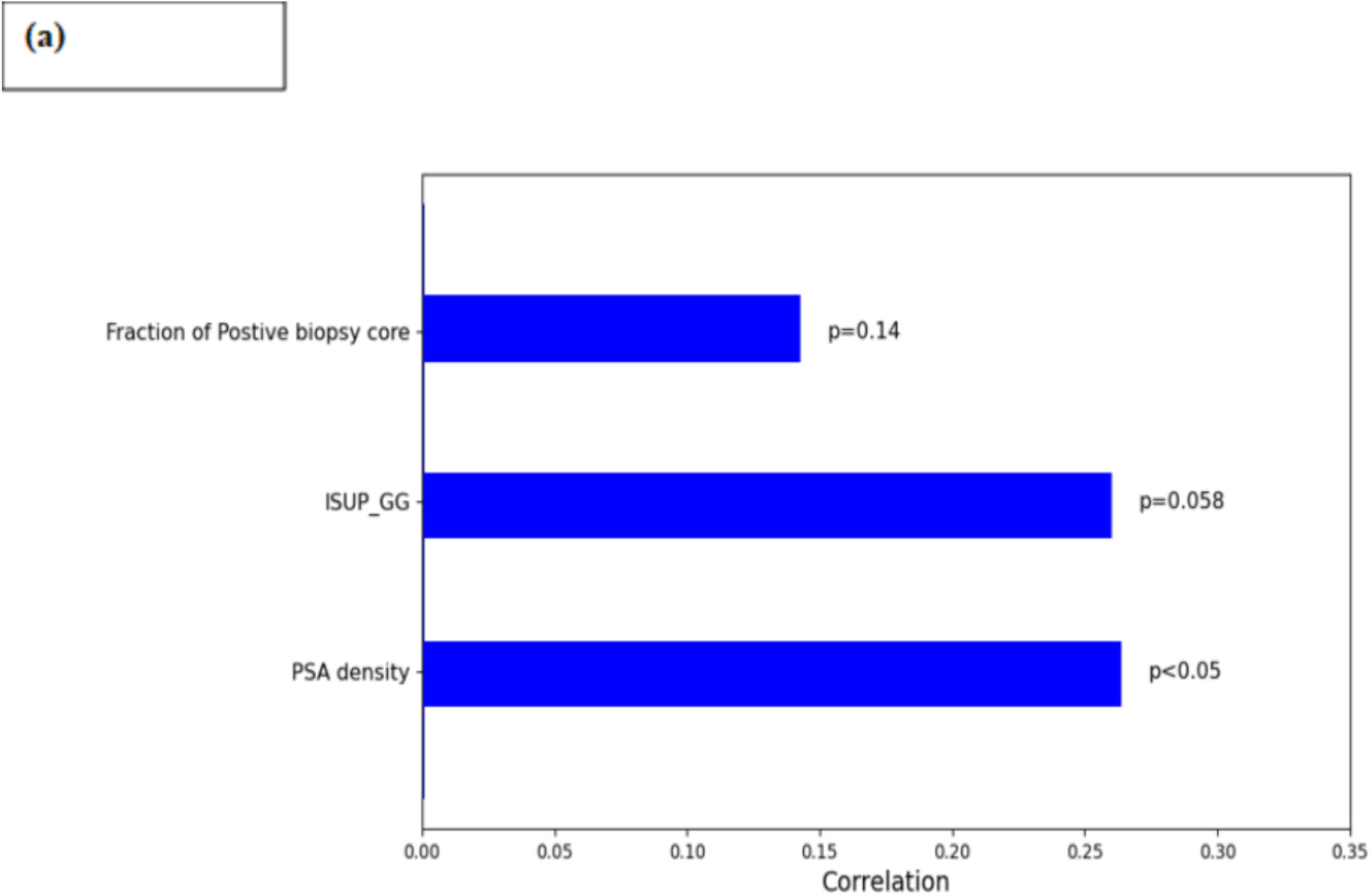
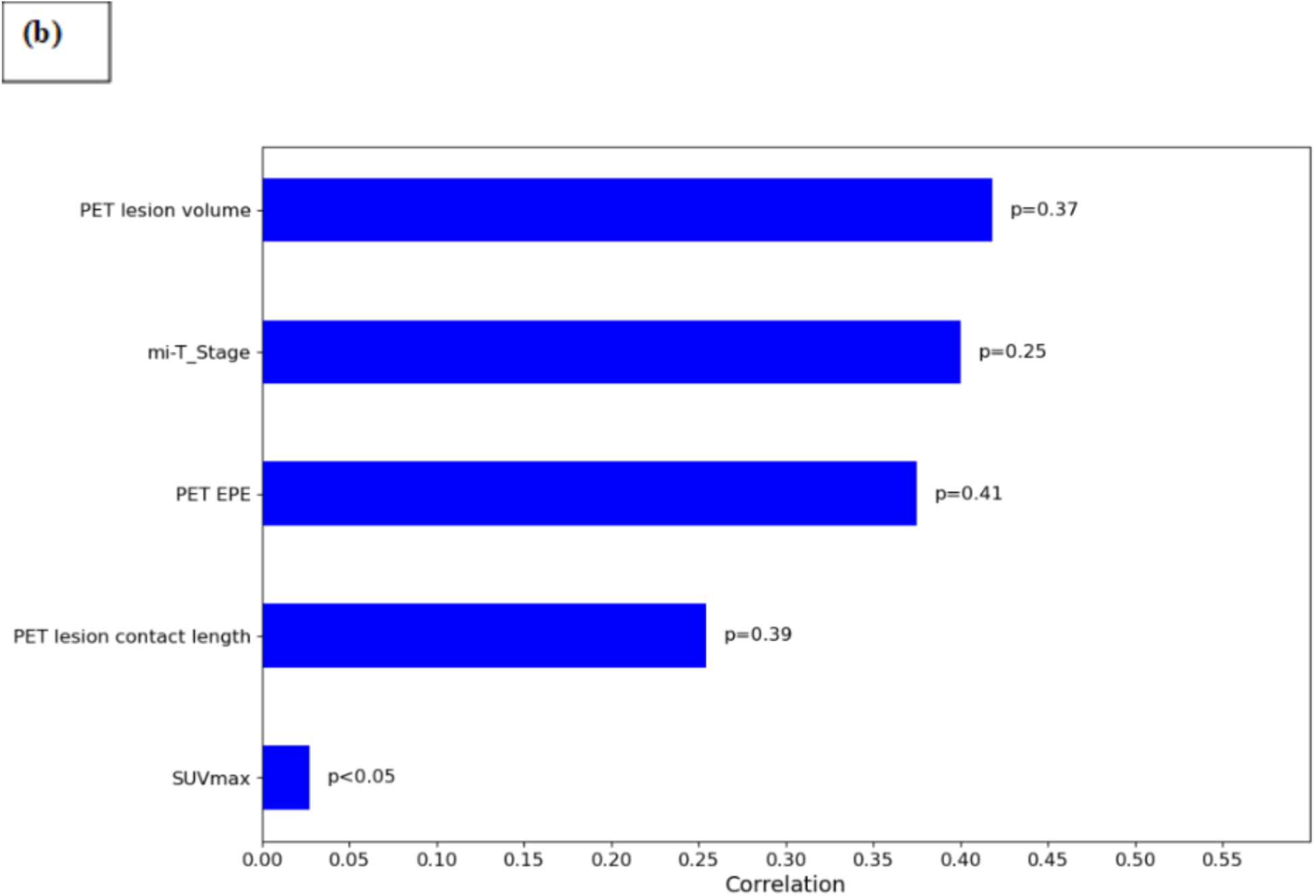
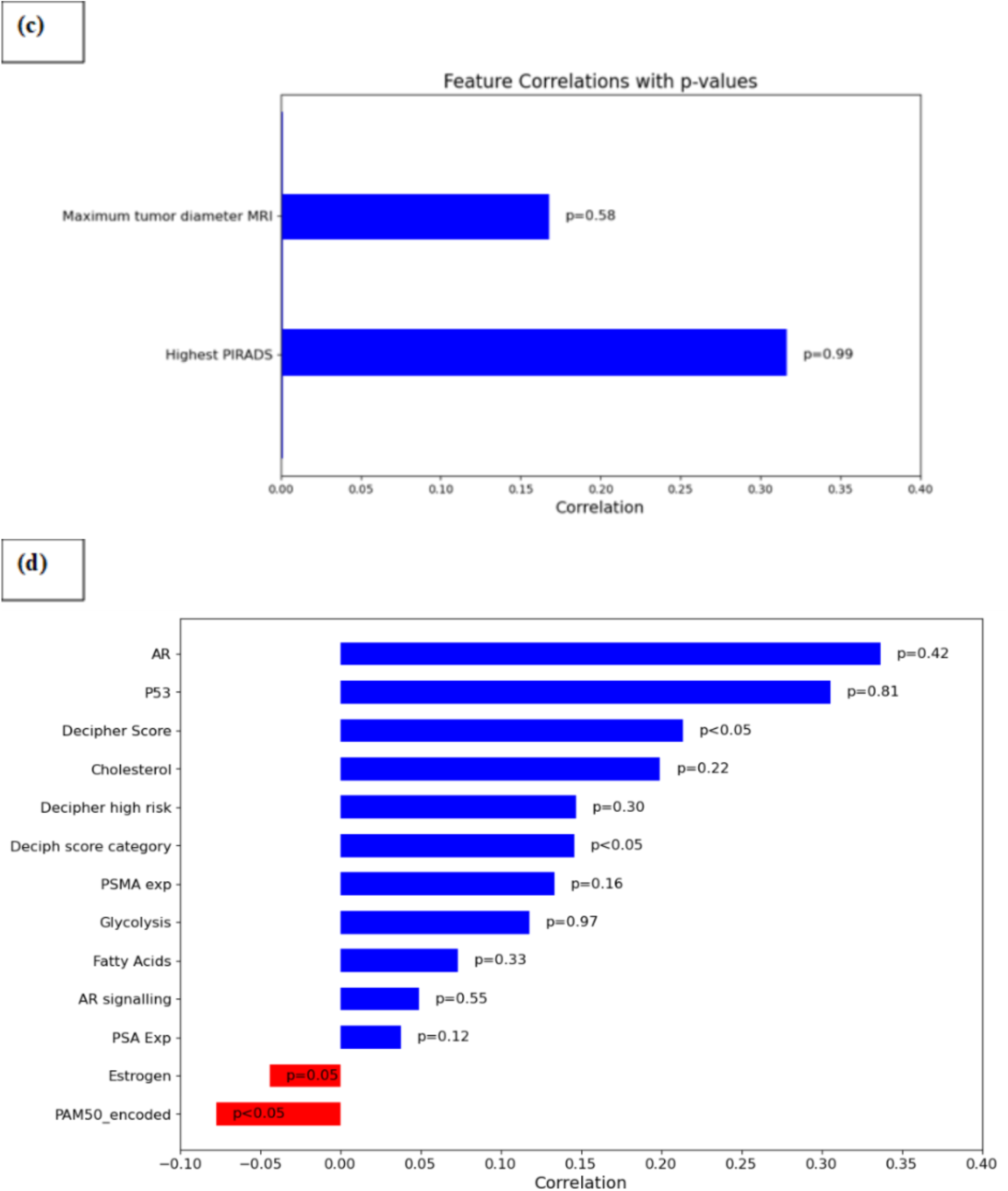
Correlation coefficients of **(a)** clinical variables: PSA density, ISUP_GG, Biopsy, PATH EPE, **(b)** PSMA-PET metrics: PET: SUV maximum, PET lesion volume, PET lesion contact length, PET EPE, **(c)** mpMRI metrics: Highest PIRADS score, maximum tumor diameter on MRI, and **(d)** Decipher Genomics: Decipher Score, Decipher score category, Decipher high risk, PAM50 (encoded), PSA Expression, PSMA Expression, AR (Androgen Receptor), P53, AR signaling, Cholesterol, Estrogen, Fatty Acids, Glycolysis.

### 2.4 Machine Learning Framework

Machine learning models were developed to predict BCR using combinations of clinical, imaging, and genomic features. Three algorithms were evaluated: Logistic Regression (LR), Random Forest (RF), and Extreme Gradient Boosting (XGBoost). Feature combinations for each model are summarized in Table 2.

**Table 2:** Characteristics of the ML-based BCR models.

| Model Types |  | Model features |
| --- | --- | --- |
| Biomarker-specific | mpMRI model | Clinical data + mpMRI |
|  | PET model | Clinical data + PSMA-PET |
|  | DGC model | Clinical data + DGC |
| Multimodal framework | PM model | Clinical data + PSMA-PET + mpMRI |
|  | PD model | Clinical data + PSMA-PET + DGC |
|  | MD model | Clinical data + DGC + mpMRI |
|  | PMD model | Clinical data + PSMA-PET + mpMRI + DGC |

LR and RF models utilized an early-fusion approach in which all selected features were combined at the input level. XGBoost models were implemented using both early- and intermediate-fusion strategies. For intermediate fusion, modality-specific features from DGC, PSMA-PET, and mpMRI were processed independently and combined prior to prediction. A localized attention mechanism was incorporated to emphasize features most relevant to recurrence prediction (Figure 2).

**Figure 2:**
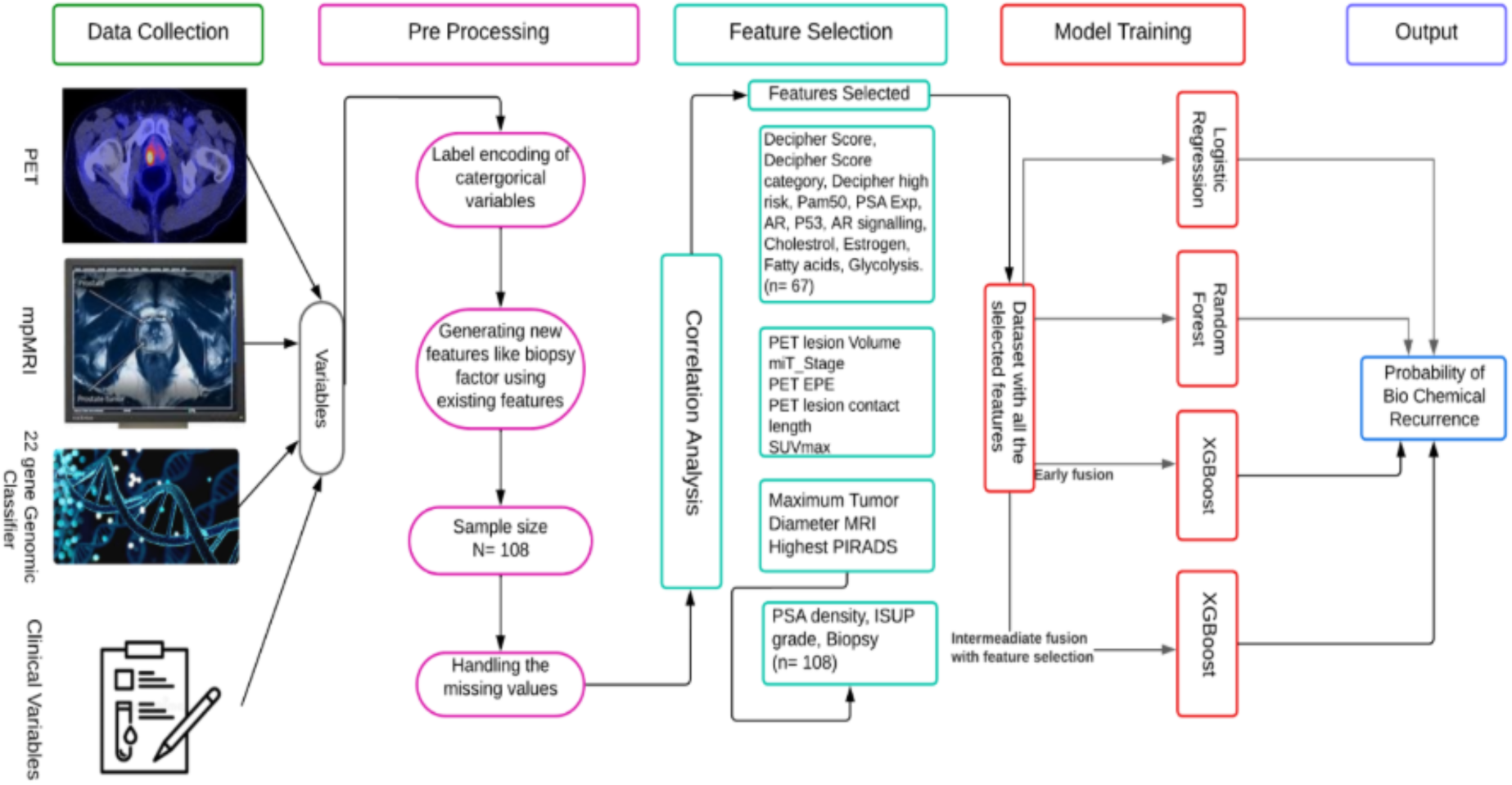
Depicts the workflow for developing the models presented in this study

Model development employed five-fold stratified cross-validation and hyperparameter optimization. Performance was evaluated using accuracy and area under the receiver operating characteristic curve (AUC).

### 2.5 Statistical Analysis and Model Evaluation

Model performance was assessed using accuracy, AUC, precision, recall, and F1-score. Decision curve analysis (DCA) was performed to evaluate clinical utility across a range of recurrence-risk thresholds by comparing model performance against treat-all and treat-none strategies.

To enhance clinical interpretability, the optimal multimodal XGBoost model was mapped onto a nomogram, utilizing a framework previously validated for extraprostatic extension prediction [18]. This nomogram generates individualized, patient-specific estimates of postoperative recurrence risk.

## 3.0 Results

The predictive performance of LR, RF, and XGBoost models was evaluated across seven feature set configurations encompassing single- and multi-modality inputs (Table 3). Across nearly all configurations, model performance ranked consistently as XGBoost Intermediate Fusion ≥ XGBoost Early Fusion > RF> LR.

**Table 3:**
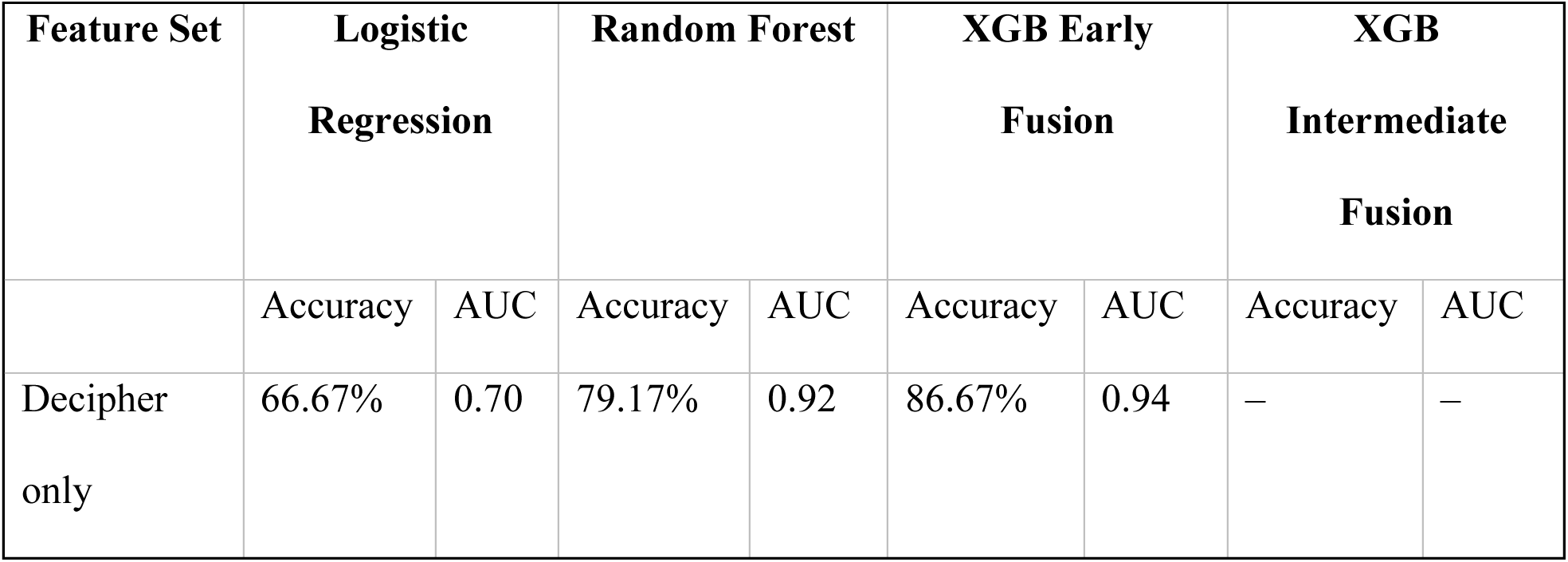
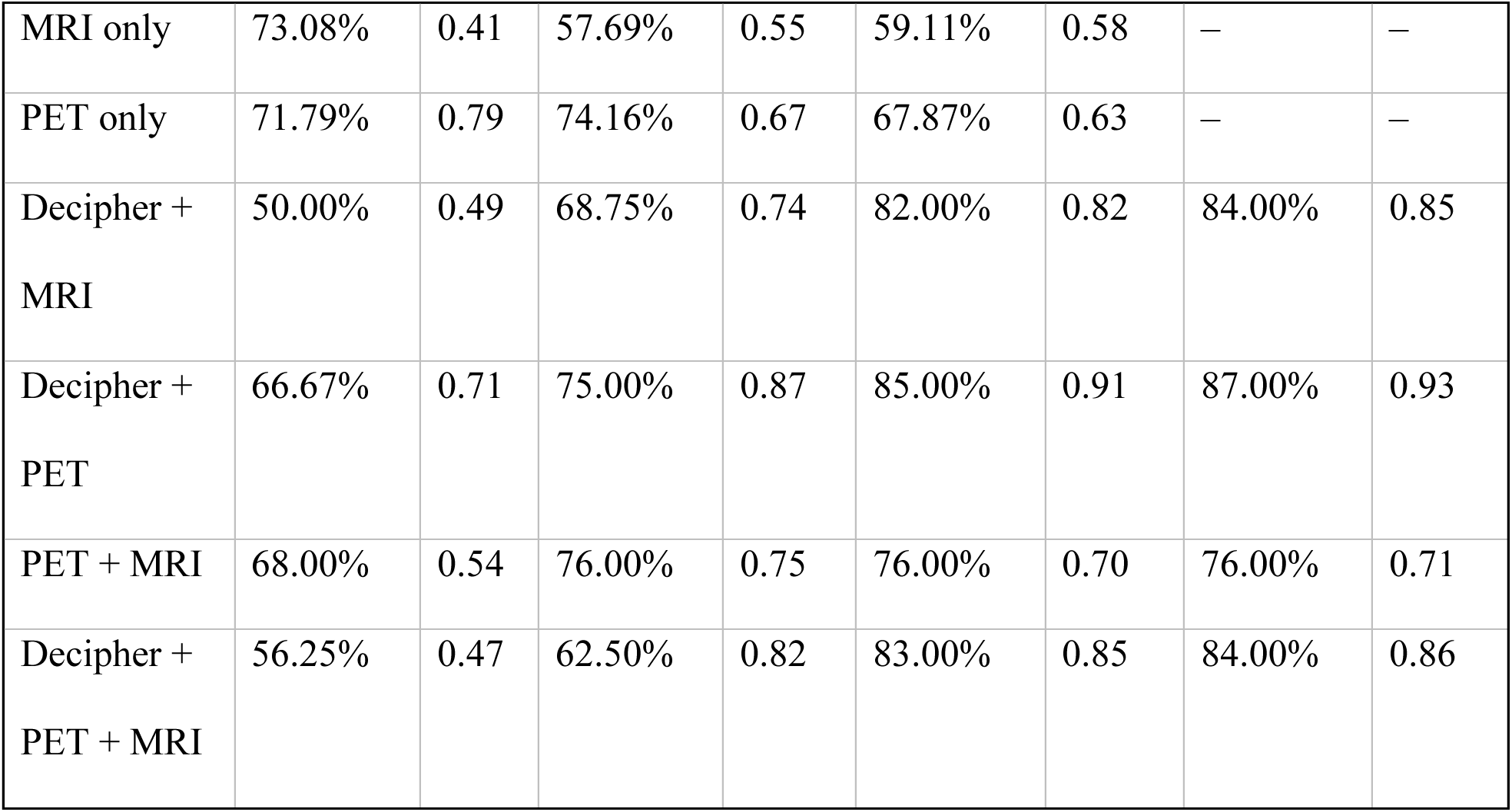
Predictive Performance of Logistic Regression, Random Forest, and XGBoost Models for Biochemical Recurrence Prediction.

Among single-modality inputs, the DGC demonstrated the strongest standalone predictive signal, with XGBoost Early Fusion achieving an AUC of 0.94 and accuracy of 86.67%. MRI-only models performed poorly across all algorithms (XGBoost AUC 0.58; logistic regression AUC 0.41), indicating limited standalone discriminative value. PET-only models showed intermediate performance, though logistic regression (AUC 0.79) and random forest (AUC 0.67) atypically outperformed XGBoost Early Fusion (AUC 0.63) in this single-modality context.

Among multi-modality combinations, the Decipher + PET feature set yielded the highest overall performance, with XGBoost Intermediate Fusion achieving an AUC of 0.93 and accuracy of 87.00%, surpassing all single-modality inputs and indicating that PSMA PET provides complementary prognostic information to Decipher genomic data. Incorporation of all three modalities (Decipher + PET + MRI) did not improve upon this two-modality combination, with XGBoost Intermediate Fusion achieving an AUC of 0.86, suggesting that MRI-derived features introduce noise or redundancy rather than incremental predictive value. The Decipher + MRI combination revealed a marked model-dependent discordance, with logistic regression performing near chance (AUC 0.49) while XGBoost models achieved AUCs of 0.82–0.85, implicating predominantly non-linear interactions between these modalities. Where applicable, XGBoost Intermediate Fusion consistently outperformed Early Fusion by 1–3 AUC points across multi-modality sets, supporting the advantage of modality-specific feature learning prior to integration.

Although the 22-gene genomic classifier demonstrated stronger independent predictive performance for BCR compared with standalone imaging modalities, PET and MRI provide complementary spatial and phenotypic characterization of tumor heterogeneity and biologic aggressiveness. Integration of these imaging-derived and molecular features through our multimodal intermediate fusion framework enabled synergistic radiogenomic modeling, where machine learning further improved prediction by leveraging complementary biologic and spatial information to enhance individualized recurrence risk stratification and predictive performance.

To facilitate individualized estimation of BCR risk and enhance the clinical interpretability, a predictive nomogram was developed based on the multimodal XGBoost intermediate fusion framework (**Figure 3**). Rather than relying on isolated variables, this model was constructed utilizing multimodal dataset, integrating Decipher genomics, PSMA-PET imaging and mpMRI features, thereby capturing a multifaceted representation of tumor biology and spatial heterogeneity to guide clinical decision-making.

**Figure 3:**
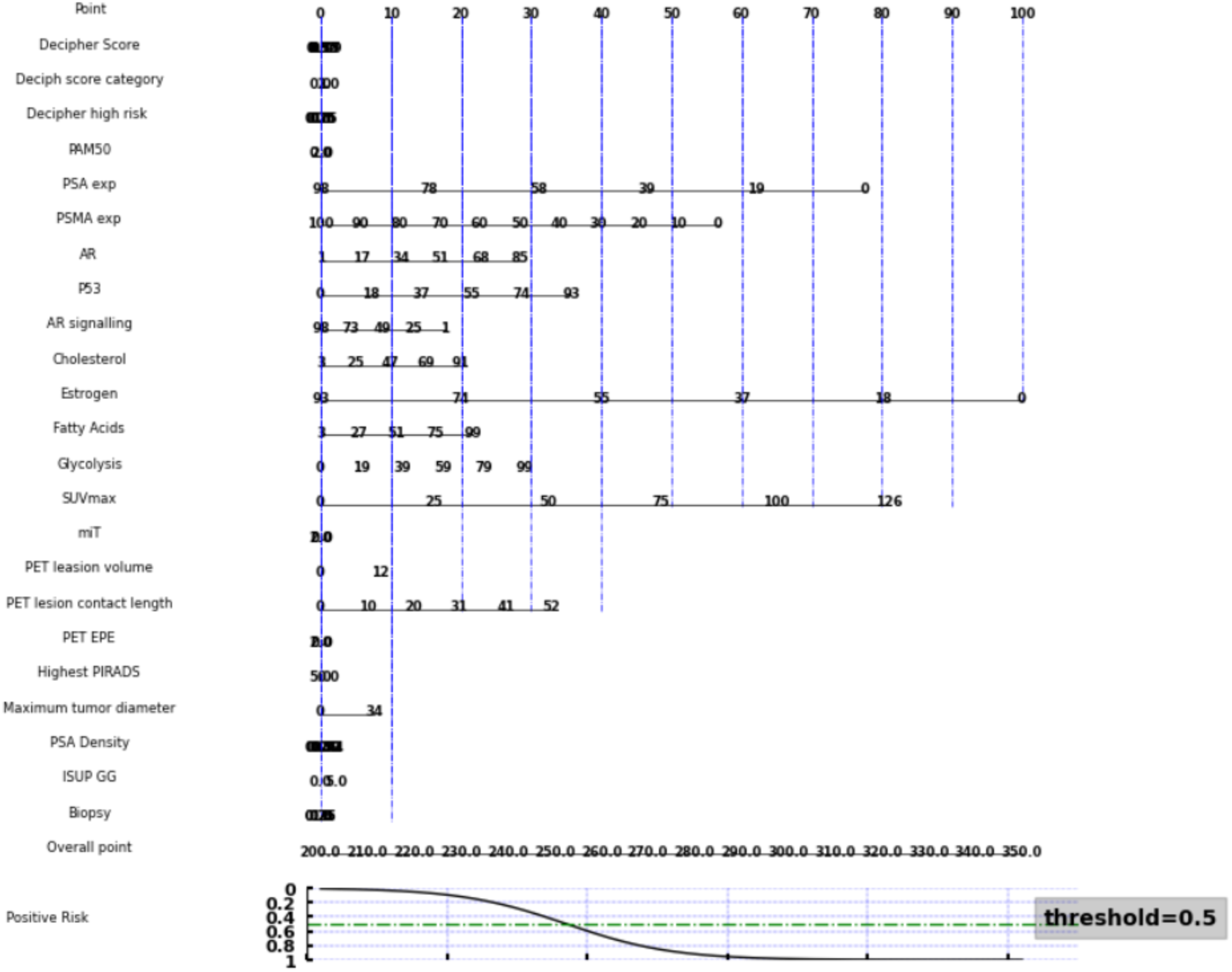
Clinical nomogram derived from the multimodal dataset (Decipher genomics, PSMA-PET, and mpMRI features) utilizing the intermediate fusion XGBoost architecture for individualized BCR prediction.

Furthermore, DCA was performed to assess the clinical net benefit of the seven XGBoost models across threshold probabilities of 0 – 1.0, with Treat All (black dashed) and Treat None (grey dash-dot) as reference strategies (**Figure 4)**. At low threshold probabilities (0–0.10), all models clustered closely together, approximating the “Treat All” line at a net benefit of approximately 0.29, with no meaningful separation between models. Divergence emerged in the clinically relevant mid-range (0.10–0.35): XGB DGC led in net benefit through approximately threshold 0.30, consistent with the dominant predictive signal of the DGC, while XGB PD exhibited the steepest early decline and fell near zero by threshold 0.30, the earliest dropout among all models. XGB PM, XGB PMD, XGB PET, and XGB MD clustered with moderate persistence through this range. Beyond threshold 0.30, where the Treat All reference crossed zero, all remaining models delivered net clinical benefit that a treat-all strategy could no longer provide.

**Figure 4:**
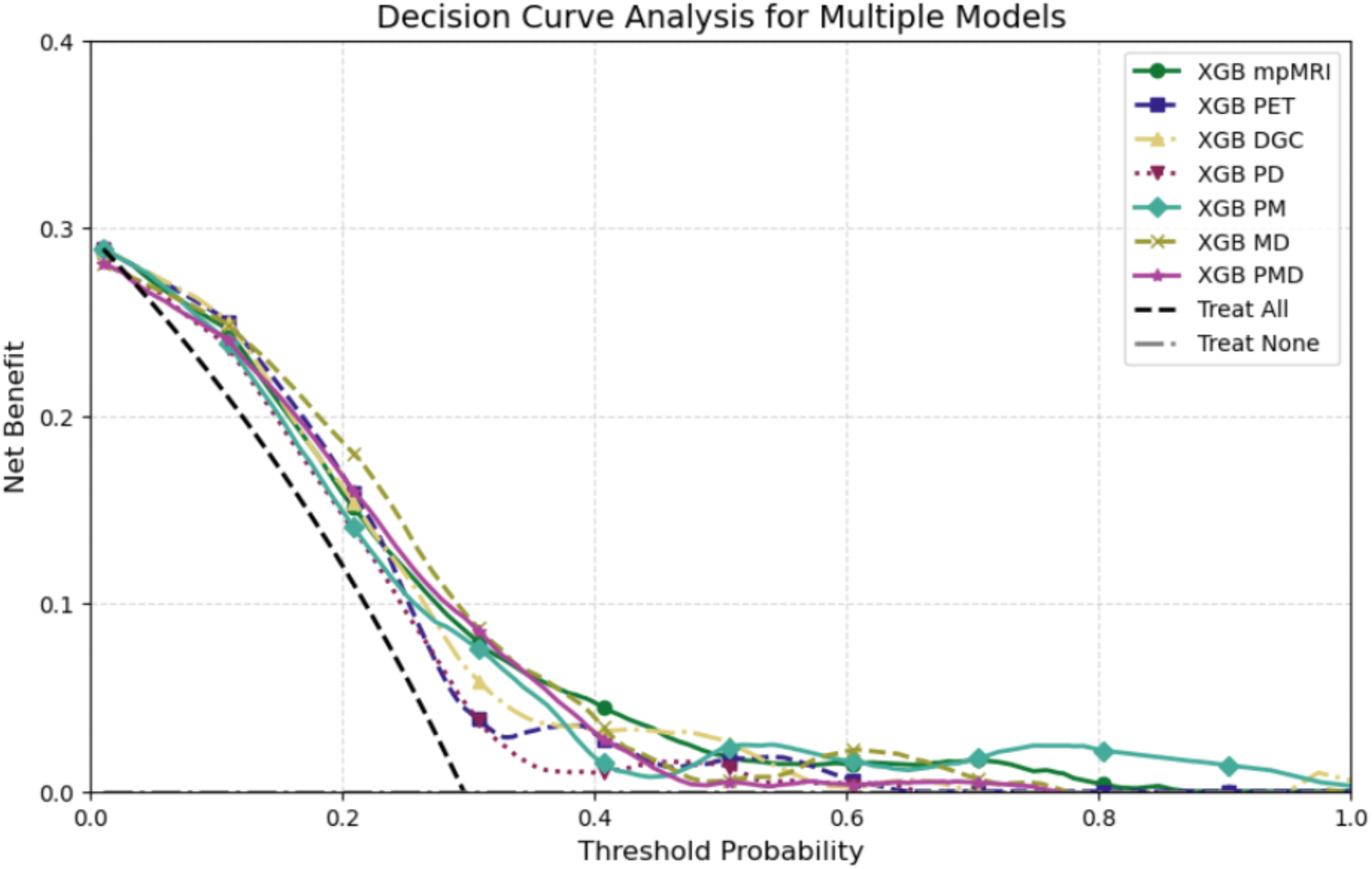
Decision Curve Analysis (DCA) demonstrates the net clinical benefit of XGBoost models across different threshold probabilities compared to “treat all” and “treat none”.

The most clinical finding was the persistence of XGB PM (PET + MRI) at high decision thresholds. While all other models converged toward zero net benefit between thresholds 0.35 and 0.50, XGB PM maintained measurable net benefit up to approximately threshold 0.80, a profile not observed in any other model, including the individual PET-only and MRI-only models. This suggests that the combination of PET and MRI captures a synergistic, highly specific signal for a subgroup of patients at elevated recurrence risk that neither modality encodes independently, and that this complementarity manifests most clearly at stricter decision thresholds. This finding is particularly notable given that XGB PET and XGB mpMRI each demonstrated only moderate-to-poor standalone AUC performance, highlighting an important dimension of multimodal integration that AUC-based analysis alone would not reveal. Critically, no model fell below zero net benefit at any threshold, confirming that all seven XGBoost models are universally non-inferior to the “treat none” strategy and would introduce no net clinical harm under any tested decision threshold.

## 4.0 Discussion

In this study, we developed and evaluated LR, RF, and XGBoost models to predict BCR following RP using multimodal data from the DGC, PSMA-PET, mpMRI, and clinical variables. Across all feature combinations, ensemble methods, particularly XGBoost with intermediate fusion and attention mechanisms, consistently outperformed LR, highlighting the importance of modeling complex nonlinear relationships across heterogeneous data sources. The strong performance of XGBoost is consistent with prior studies demonstrating the utility of gradient-boosting approaches for prostate cancer outcome prediction [22].

Among the evaluated modalities, DGC emerged as the strongest individual predictor, achieving an AUC of 0.94 in the XGBoost framework. This finding aligns with extensive evidence supporting the prognostic value of the Decipher classifier for recurrence, metastasis, and prostate cancer-specific mortality [15–17]. While previous studies have largely evaluated DGC as a standalone biomarker or in combination with clinical nomograms, our results suggest that machine learning-based integration with imaging biomarkers may further enhance predictive performance.

PSMA-PET demonstrated moderate standalone predictive value but provided complementary information when combined with DGC. This observation is consistent with previous reports showing that PET-derived biomarkers, including SUVmax, tumor burden, and nodal involvement, are associated with recurrence risk [13,14,23]. The improved performance observed in multimodal models suggests that DGC and PSMA-PET capture distinct aspects of tumor biology, with genomics reflecting intrinsic tumor aggressiveness and PET characterizing disease burden and spatial dissemination.

In contrast, MRI contributed less predictive value than genomic and PET biomarkers. Previous studies using quantitative MRI radiomic or deep-learning features have reported improved BCR prediction [24,25]. However, our analysis relied on clinically reported PI-RADS scores rather than high-dimensional imaging features, which may explain the lower performance observed. Although PI-RADS provides a standardized and clinically interpretable assessment, it may not fully capture the underlying tumor heterogeneity present within mpMRI data.

A notable finding of this study was the consistent advantage of intermediate fusion over conventional early fusion. By preserving modality-specific information before integration, intermediate fusion may better capture cross-modal interactions while reducing feature redundancy. This observation is consistent with emerging multimodal oncology frameworks that have demonstrated improved performance using intermediate fusion strategies [26,27]. Our results extend these findings to structured genomic, imaging, and clinical data, demonstrating that attention-weighted multimodal integration can improve BCR prediction without requiring complex deep-learning architectures.

Comparing our model to the EAU biochemical recurrence risk stratification framework [28]. While EAU risk groups provide clinically validated categorical stratification based on clinicopathologic variables, they lack granularity at the individual patient level. Our multimodal machine learning model incorporates genomic, imaging, and molecular biomarkers to provide continuous patient-specific risk estimates, enabling finer stratification within conventional EAU risk categories.

To our knowledge, this is among the first studies to systematically evaluate DGC, PSMA-PET, and mpMRI within a unified machine learning framework for preoperative BCR prediction following radical prostatectomy. Previous studies have primarily focused on individual modalities or pairwise combinations, whereas our analysis directly compared the relative and combined contributions of genomic, molecular imaging, and radiologic biomarkers. The findings support the value of multimodal integration for improving personalized risk assessment.

The proposed multimodal framework has the potential to improve preoperative risk stratification by integrating complementary genomic and imaging biomarkers into a single predictive model. More accurate identification of patients at high risk of biochemical recurrence could facilitate individualized patient counseling, inform treatment planning, and support decisions regarding surgical management, multimodal therapy, and postoperative surveillance. Following external validation in larger prospective cohorts, such models may serve as clinically useful decision-support tools to complement existing risk stratification approaches.

Several limitations should be acknowledged. First, the retrospective single-institution design and modest sample size may represent a selected population with a higher pretest risk of recurrence, which may limit model generalizability. Second, the study focused on binary recurrence outcomes rather than time-to-event analysis. Third, MRI assessment was based on clinically reported PI-RADS scores rather than standardized radiomic features, potentially limiting the prognostic information captured. Finally, external validation was not performed and will be necessary before clinical implementation.

Future work should focus on validating these findings in larger multi-institutional cohorts, incorporating standardized radiomic pipelines, and exploring survival-based machine learning approaches capable of modeling recurrence timing. Such efforts may further improve the robustness and clinical applicability of multimodal radiogenomic prediction models.

## 5. Conclusion

This study demonstrates that integrating DGC, PSMA-PET, and mpMRI biomarkers using machine learning improves prediction of biochemical recurrence following radical prostatectomy. XGBoost with intermediate fusion achieved the strongest overall performance, with the combination of DGC and PSMA-PET providing the greatest predictive value. DGC emerged as the most informative individual modality, while PSMA-PET contributed complementary prognostic information that enhanced risk stratification.

The superior performance of intermediate fusion, favorable decision curve analysis results, and successful development of an interpretable nomogram highlight the potential clinical utility of multimodal machine learning for individualized postoperative risk assessment. Future studies should focus on external validation, incorporation of quantitative imaging biomarkers, and survival-based modeling approaches to further improve clinical translation and generalizability.

## Declarations

### Ethics approval and consent to participate

This retrospective study was approved by the Indiana University Institutional Review Board (IRB No: 13892). The requirement for informed consent was waived due to the retrospective nature of the study and the use of de-identified clinical data.

### Consent for publication

Not applicable.

### Availability of data and materials

The datasets analyzed during the current study are not publicly available because they contain protected patient information and are subject to institutional data-sharing restrictions. De-identified data may be made available from the corresponding author upon reasonable request and with permission from Indiana University.

### Authors’ contributions

R.C conceived the study, curated the data, performed the statistical and machine learning analyses, interpreted the results, and drafted the manuscript. O.O conceived and supervised the study, provided clinical guidance, interpreted the findings, and critically revised the manuscript. C.Y, H. L, R.S, and B.N contributed to data acquisition, clinical interpretation, and critical revision of the manuscript. All authors read and approved the final manuscript.

## Data Availability

De-identified data is available upon request.

## Acknowledgements

The authors thank all collaborators and clinical staff who contributed to patient care and data collection for this study.

